# Neuroplastic Effects of Dance Training in Parkinson’s Disease: Functional and Structural Modulation in Speech-Related Brain Regions

**DOI:** 10.1101/2025.07.03.25330757

**Authors:** Ashkan Karimi, Xianze Meng, Karolina A. Bearss, Sarah Robichaud, Rachel J. Bar, Joseph F.X. DeSouza

## Abstract

**Background:** Parkinson’s disease (PD) is a neurodegenerative disorder characterized by progressive motor and non-motor impairments, including speech deficits. Dance-based interventions have been proposed as a promising rehabilitative strategy for enhancing motor function, cognitive engagement, and neuroplasticity. However, the neural mechanisms underlying dance-induced neuroplasticity remain poorly understood.

**Objective:** This study aimed to investigate the effects of an 8-month dance training program on functional brain activity, cortical structure, and white matter integrity in people with PD (PwPD) using a multi-modal neuroimaging approach.

**Methods:** Ten individuals with PD participated in dance training and underwent functional MRI (fMRI) and cortical thickness analysis (T1-weighted MRI) at four time points, and 4 of them underwent diffusion-weighted imaging (DWI) at the last two time points. fMRI examined BOLD signal changes in the motor cortex (inferior frontal gyrus) and the supplementary motor area (SMA) during a dance-imagery task. Cortical thickness was measured in Broca’s area and the left supplementary frontal language (SFL) area. A reference group from the Michael J. Fox Foundation’s Parkinson’s Progression Markers Initiative (PPMI) dataset was selected for cortical thickness analysis. White matter integrity between Broca’s area and the SFL area was assessed using fractional anisotropy (FA), axial diffusivity (AD), radial diffusivity (RD), and mean diffusivity (MD). Additionally, fundamental frequency standard deviation (F0SD), a key indicator of speech prosody, was analyzed in three participants over a 5-year follow-up period to examine the functional relevance of neuroplastic changes.

**Results:** Reductions in BOLD signal were observed in the motor cortex (inferior frontal gyrus) between September and January (*p* = 0.021), suggesting increased neural efficiency. In contrast, SMA exhibited a significant increase in BOLD signal activity between September and January (*p* = 0.010), indicating enhanced motor-speech integration. No further notable changes in functional activity in both regions were detected between January and April. The dance group showed significant changes in cortical thickness in the left SFL area across time, with increases from September to December (*p* = 0.049) followed by decreases from December to April (*p* = 0.046). However, no changes were detected in Broca’s area for the dance or reference groups. Additionally, the reference group did not exhibit any significant changes in the left SFL area across the same time points. FA increased significantly between January and April (*p* = 0.024), suggesting enhanced white matter organization in speech-motor pathways. AD showed a marginal increase during this period (*p* = 0.056), while RD and MD remained stable. Critically, voice feature analysis revealed significant improvements in F0SD over the 5-year follow-up period (*p* < 0.0001), demonstrating sustained functional benefits in speech prosody that were associated with the earlier neuroplastic adaptations.

**Conclusion:** Dance training was associated with significant functional and white matter neuroplastic changes in PD, particularly in motor-related functional activity and white matter connectivity between motor and language networks. The neuroplastic adaptations observed during the 8-month intervention period were associated with sustained improvements in speech prosody that persisted for years, providing compelling evidence for the clinical relevance of dance-induced brain changes. While dynamic cortical thickness changes were observed in the SFL area, the functional and structural connectivity adaptations appear to be the primary drivers of therapeutic benefit. These findings highlight dance as a promising, intervention for enhancing neural efficiency and motor-linguistic integration in PD, with demonstrated long-term functional benefits that warrant further investigation in larger, controlled studies.

## 1. INTRODUCTION

### 1.1. Parkinson’s Disease and Its Neural Implications

Parkinson’s Disease (PD) is a progressive, age-related neurodegenerative disorder marked by both motor and non-motor symptoms^1^. It primarily arises from the degeneration of dopamine-producing neurons in the substantia nigra pars compacta^2^. The prevalence of PD increases with age, affecting around 1% of individuals aged 65–69, and rising to nearly 3% in those aged 80 and older^3^. Due to its progressive nature, effective interventions must address both motor and non-motor symptoms to help maintain the quality of life (QoL) in people living with PD (PwPD).

In addition to motor impairments, PD is strongly associated with speech difficulties, affecting up to 90% of individuals^4^. These often present as hypokinetic dysarthria, characterized by reduced pitch variation, diminished vocal intensity, and monotonous speech. Speech impairments can appear in the early stages of the disease and tend to worsen over time, significantly hindering communication and social interaction, hence decreasing the QoL for both PwPD and their caregivers. Although pharmacological treatments like levodopa, dopamine agonists, and Monoamine Oxidase-B (MAO-B) inhibitors can alleviate symptoms^5^, they do not slow disease progression and may produce notable side effects^6,7^. The side effects include, but are not limited to, fibrosis, sleep attacks, impulse control disorders, depression, hypotension, and liver toxicity^8^. Deep brain stimulation (DBS) has also emerged as a surgical intervention for managing motor symptoms in advanced PD. While DBS can significantly improve motor function and reduce medication requirements, its long-term efficacy remains controversial. Some studies highlight sustained benefits over time, whereas others report a decline in therapeutic effects, along with potential complications such as worsening of speech, gait, cognition, and postural stability^9,10^. Additionally, patient satisfaction with DBS outcomes appears to vary considerably, often influenced by disease progression and unmet expectations^11^.

As a result, there is increasing interest in complementary therapies, such as physical exercise, music therapy, and dance, that aim to enhance neuroplasticity and support functional recovery^12–17^.

### 1.2. Dance as a Neuroplasticity-Based Intervention in PD

Emerging research suggests that engaging in physical and cognitive activities, such as dance, may offer neuroprotective benefits for PwPD^18–22^. Dance-based interventions stimulate multiple brain networks involved in motor control, coordination, rhythm processing, and executive functioning, providing therapeutic effects that may extend beyond those of conventional rehabilitation approaches. Neuroimaging studies have demonstrated that dance may enhance neuroplasticity in key motor regions, including the supplementary motor area (SMA), putamen, and caudate nucleus within the basal ganglia^23^. These neural adaptations may support improvements in motor performance, balance, and gait and positively influence cognitive and emotional well-being in PwPD^18–20^.

While prior studies have examined the motor and functional outcomes of dance interventions in PD, their influence on neuroplastic changes remains less well understood. In particular, it is still unclear how dance training may drive structural and functional changes within the brain’s motor and language networks. Since speech impairments in PD are believed to result from dysfunction across both cortical and subcortical regions, investigating how dance affects brain areas associated with word production could yield valuable insights into its therapeutic potential for addressing speech and motor coordination deficits.

Building upon this, our previous study demonstrated that weekly dance training in PwPD led to significant improvements in vocal performance, particularly in measures of fundamental frequency standard deviation (F0SD), a critical indicator of prosodic variability^22^. While F0SD does decline as PD progresses^24,25^, our findings suggested that dance-based interventions preserved speech prosody in 29 PwPD, reflecting possible engagement and plasticity within speech-motor integration pathways.

Improvements in F0SD could be associated with more robust activation of cortico-subcortical circuits, including Broca’s area, the SMA, and their connections with subcortical structures such as the basal ganglia and cerebellum, which are essential for precise vocal control. This highlights the importance of examining dance-induced changes in both functional activation and structural connectivity within speech-motor networks. By linking behavioral improvements in speech for PwPD (blue bars) who dance over 5-yrs (Karimi et al., 2025, **FIGURE 1**) with neuroimaging markers of brain plasticity, we can deepen our understanding of how dance interventions promote adaptive reorganization across multiple domains affected by PD.

**FIGURE 1.**
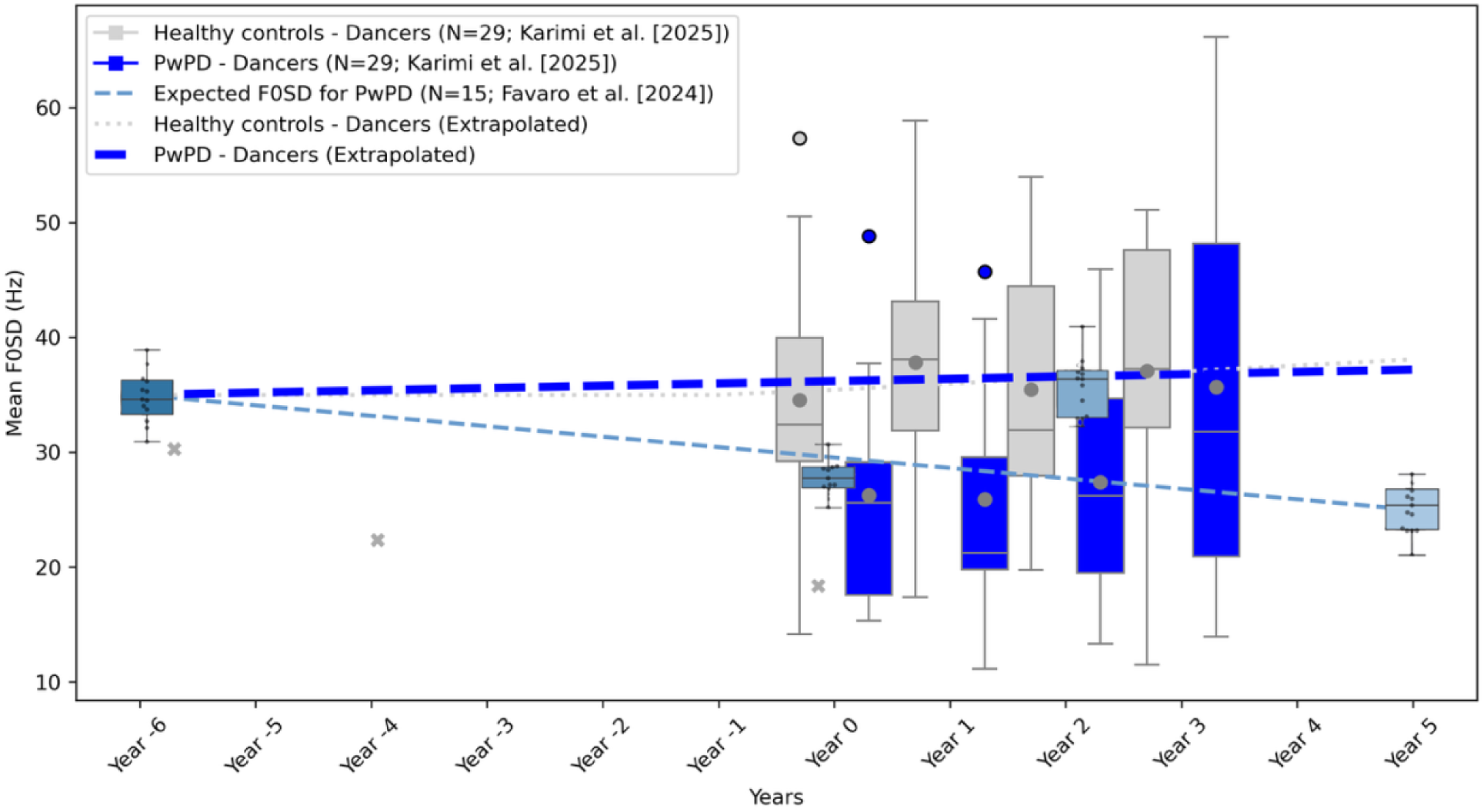
illustrates F0SD trajectories for PwPD who dance (dark blue) and do not dance (light blue). PwPD dancers from our longitudinal behavioural speech study (2014 to 2019) in comparison to PwPD nondancers from Favaro et al. (2024) and Harel et al. (2004) [✖]. These behavioral findings provide further rationale for examining dance-induced changes in speech-motor brain regions using multimodal imaging approaches.

### 1.3. Cortical Thickness and White Matter Connectivity as Biomarkers of Neuroplasticity

Neuroplasticity—also referred to as brain plasticity or neural plasticity—is the brain’s capacity to undergo adaptive structural and functional changes^26^. Mateos-Aparicio & Rodrigues- Moreno^27^ defined neuroplasticity as “the ability of the nervous system to change its activity in response to intrinsic or extrinsic stimuli by reorganizing its structure, functions, or connections.”.

#### 1.3.1 Cortical Thickness as an Indicator of Structural Adaptation

Cortical thickness, measured using T1-weighted structural MRI, reflects the morphological integrity of the cerebral cortex. Changes in cortical thickness have been linked to learning, motor training, and rehabilitation, making it a valuable biomarker for neuroplasticity^28^. Studies have shown that motor and cognitive training can lead to increases in cortical thickness, particularly in motor and prefrontal regions associated with skill acquisition and executive control^29,30^.

Conversely, cortical thinning in PD has been associated with disease progression, cognitive impairment, and motor decline^31^. Assessing cortical thickness in speech and motor-related regions, such as Broca’s area and the left supplementary frontal language (SFL) area, may reveal structural adaptations associated with dance training in PD.

#### 1.3.2 DTI/DWI and White Matter Plasticity

Among the imaging methods used to study structural neuroplasticity, diffusion MRI - particularly diffusion tensor imaging (DTI)- is considered a powerful tool for probing microstructural properties of brain tissue^32^. Unlike conventional anatomical MRI, DTI provides more specific insights into plasticity-related morphological changes. It offers various indices, such as fractional anisotropy (FA), which is commonly used to assess the integrity and organization of white matter fibers^33^. Other metrics, such as Mean Diffusivity (MD), Axial Diffusivity (AD), and Radial Diffusivity (RD), provide insights into different aspects of microstructural changes^34^. Several studies have reported significant training-induced alterations in DTI parameters within relevant white matter pathways, suggesting that DTI can potentially detect structural plasticity in both gray and white matter^35–37^. Nonetheless, these changes’ precise biological and morphological interpretations are still not fully understood^38^.

Motor learning, language acquisition, and rehabilitation studies have reported changes in white matter connectivity following training interventions^39,40^. In PD, progressive white matter degeneration affects motor and speech networks, leading to communication deficits and impaired movement execution^41^. Examining structural connectivity changes between Broca’s area and the left SFL area using DWI/DTI can provide insights into whether dance training enhances white matter integrity in language-motor pathways.

### 1.4. Motivation and Study Objectives

The motivation for this study stemmed from our findings in a recent investigation of voice features in 29 dancers with PD, which demonstrated that dance training significantly improved the standard deviation of fundamental frequency (F0SD) in PwPD^22^. This study highlighted the potential of dance as a therapeutic intervention for addressing speech impairments in PD.

Building on these results, the current study aimed to explore the neural mechanisms underlying such improvements by examining the neuroplastic effects of dance using multi-modal neuroimaging techniques. Specifically, we examined:

- Functional activity in the motor cortex (inferior frontal gyrus) and the supplementary motor area (SMA) using functional MRI (fMRI).
- Cortical thickness in Broca’s area and the left supplementary frontal language (SFL) area using structural MRI analysis.
- White matter integrity between Broca’s and left SFL areas using DWI.

The multi-model MRI data (functional, structural, and diffusion) were collected between September 2013 and April 2014 as part of the initial phase of the study^18,19,21^. Following this, from 2014 until the COVID-19 pandemic, a longitudinal follow-up study was conducted to examine motor and non-motor symptoms in PwPD who attended weekly dance classes^20^. While F0SD measures were obtained during and after the MRI data acquisition, they offer a valuable opportunity to relate neuroimaging findings to later observed changes in voice features.

For three PwPD who participated in both studies, we analyzed their F0SD trajectories over the follow-up period to examine potential associations between early neuroimaging markers and longitudinal changes in speech characteristics. The F0SD trajectories for these individuals, visualized in **FIGURE 3**, illustrate both the within-subject variability and the overall pattern of change across pre- and post-dance session assessments. These retrospective observations provide an important extension of the original voice feature study, linking longitudinal behavioral improvements with neuroimaging data.

We hypothesized that dance training would lead to functional adaptations in motor regions, structural changes in cortical thickness, and enhanced white matter connectivity between motor and language networks. By integrating fMRI, cortical thickness analysis, and DWI, this study provides a comprehensive investigation of dance-induced neuroplasticity in PD, with potential implications for rehabilitation strategies and non-pharmacological interventions.

## 2. METHODS

### 2.1. Overview and Participants

Participants were recruited from weekly, 75-minute, community-based Sharing Dance Parkinson’s (SDPD) classes at Canada’s National Ballet School ([NBS](https://www.nbs-enb.ca/dance-for-life/sharing-dance-parkinson-s/)). Recruitment occurred at the start of a dance term, participation was voluntary, and interested individuals received written information before deciding to participate. Participants’ characteristics are summarized in TABLE 1. Participants self-reported their health status. All PwPD were on stable medication regimens throughout the study period.

**TABLE 1.**
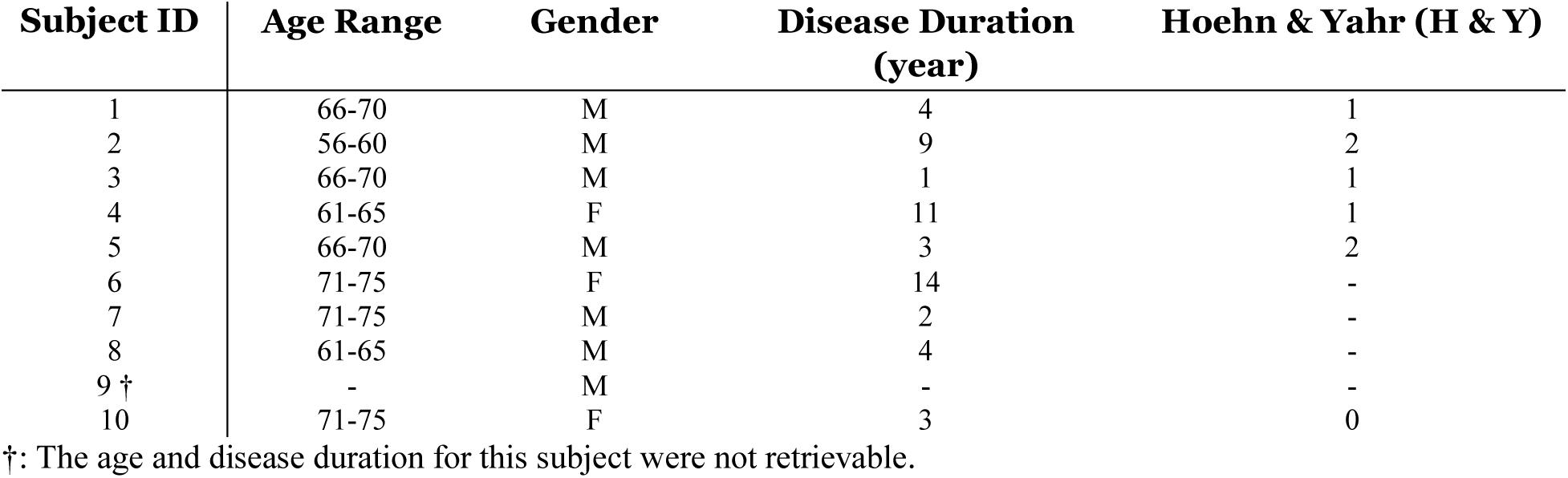
Characteristics of People with Parkinson’s Disease.

| Subject ID | Age Range | Gender | Disease Duration<br>(year) | Hoehn & Yahr (H & Y) |
| --- | --- | --- | --- | --- |
| 1 | 66-70 | M | 4 | 1 |
| 2 | 56-60 | M | 9 | 2 |
| 3 | 66-70 | M | 1 | 1 |
| 4 | 61-65 | F | 11 | 1 |
| 5 | 66-70 | M | 3 | 2 |
| 6 | 71-75 | F | 14 | - |
| 7 | 71-75 | M | 2 | - |
| 8 | 61-65 | M | 4 | - |
| 9 † | - | M | - | - |
| 10 | 71-75 | F | 3 | 0 |

†: The age and disease duration for this subject were not retrievable.

SDPD classes are based on the Mark Morris Dance Group’s Dance for PD® protocol^42^ and follow a structured format, progressing from seated exercises to paired and group movements, culminating in standing choreography and locomotion through space. The study’s choreography emphasized social interaction, with participants learning and practicing sequences weekly from September to June, leading to three public performances: Sharing Dance Day, Toronto City Hall (for Parkisnon’s Awareness Month), and the Parkinson’s Central Annual Meeting. For dance protocol refer to Bearss et al. (2017) study^19^.

### 2.2. MRI Data Acquisition

MRI data were acquired using a 3T Siemens Tim Trio scanner with a 32-channel head coil at York University’s Sherman Health Sciences Centre. Functional and structural data were collected across four time points (September 2013, December 2013, January 2014, April 2014) over an 8-month period from ten participants. A subset of four participants underwent DWI in January and April scans. The imaging modalities included fMRI for functional activity, T1- weighted scans for cortical thickness, and DWI for white matter connectivity. This multi-modal imaging approach provided insights into functional activation, structural changes, and connectivity adaptations in response to dance training in PwPD.

### 2.3. fMRI Analysis

#### 2.3.1. fMRI Task Paradigm

Participants performed an imagery-based dance task while in the scanner. They were instructed to mentally rehearse the choreography practiced during dance classes from a first-person/internal perspective, engaging both visual and kinesthetic modalities. The first minute of the associated music was played through headphones to facilitate accurately timed imagery.

A block design was employed, alternating between ON state (dance-imagery task)-60 seconds of imagined dance movement, and OFF state (fixation block)-30 seconds of resting fixation (FIGURE 2).

**FIGURE 2.**
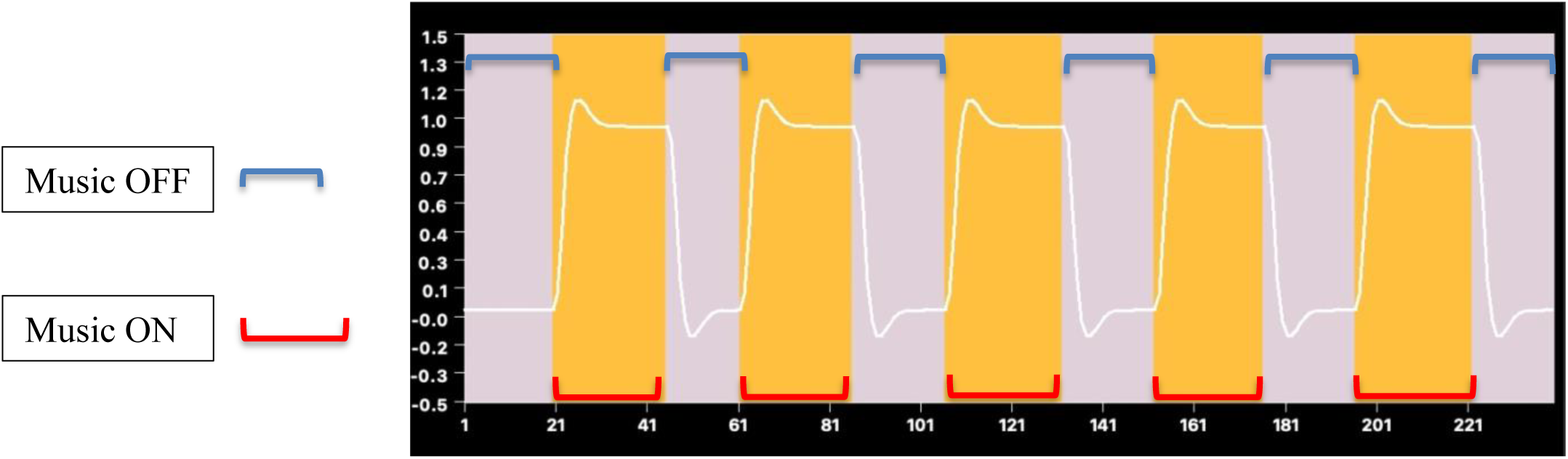
Block design of music ON (red) and music OFF (blue).

Each participant completed five ON/OFF cycles, with a total scan duration of 8 minutes. The first 15 volumes were discarded from the analysis to allow for magnetic field stabilization.

#### 2.3.2. fMRI Preprocessing

Preprocessing was conducted using BrainVoyager QX version 22.0.3.4578, following standard pipelines. Functional data were superimposed on an anatomical scan and transformed into Talairach space. The preprocessing steps included slice timing correction – temporal alignment of slices within each volume, motion correction – realignment of functional images; maximum displacement did not exceed 1 mm (translation) or 3 mm (rotation) across time points, temporal high-pass filtering – Fourier-based filtering to remove low-frequency drifts and physiological noise, using a General Linear Model (GLM) incorporating two sine and two cosine functions, spatial normalization – transformation of functional data into Talairach space for group-level analysis, and artifact removal – no scans were excluded, as all motion correction values were within acceptable limits.

#### 2.3.3. Region of Interest (ROI) Selection

ROIs were selected based on their relevance to motor and speech functions: ***Motor cortex (inferior frontal gyrus).*** ROI mask was defined using BrainVoyager’s built-in ROI tools and centered at the Talairach coordinates (*x* = -65, *y* = 6, *z* = 18) with a 4-mm radius (257 voxels)^43^. ***SMA.*** An additional ROI was selected in the SMA based on prior literature identifying SMA regions associated with vocalization^44^. Specifically, coordinates reported at (x = -3.1, y = 9.4, z = 52) were transformed into Talairach space^45^, resulting in a final center at (x = -2, y = 11, z = 47). A spherical ROI with a radius of 10.1 mm, encompassing 4385 voxels, was defined around this center point^21^. This SMA ROI was included due to its involvement in both motor planning and speech production processes.

Given that the dance choreography included a vocalization component—participants simulated holding a lasso and vocalized “yeeeee haw” between the 20th and 30th seconds of the routine—functional MRI signals were specifically extracted from this time window during the imagery task. BOLD signal changes within the SMA ROI during this vocalization imagery period were analyzed to assess neuroplastic adaptations related to integrated motor-speech processing.

#### 2.3.4. fMRI Analysis

A fixed-effects multi-subject analysis was conducted to compare brain activity during dance-imagery blocks and fixation blocks at each time point. Functionally defined regions were identified from the GLM contrast (dance-imagery vs. fixation) across all time points, with a statistical threshold of *p* < 0.0001 (Bonferroni-corrected) and a cluster threshold of k > 22.

BOLD percent signal change was calculated relative to a baseline, which is defined as the average of the two volumes acquired before the music onset.

To examine longitudinal changes in task-related BOLD modulation, linear mixed-effects models were used, with time as a fixed factor, with the baseline as the reference level. Random effects for individual samples to account for inter-subject variability. Autoregressive correlation structure of order 1 (AR [1]) to model correlations between consecutive samples.

### 2.4. Cortical Thickness Analysis

#### 2.4.1 Cortical Thickness Extraction

Cortical thickness was measured from T1-weighted anatomical scans using FreeSurfer version 7.4.1^46^. The “recon-all” processing pipeline was applied to each T1-weighted scan for each subject to extract cortical surface measurements automatically.

#### 2.4.2 ROI Selection

ROIs were defined using the Human Connectome Map^47^ and selected based on their proximity to the fMRI ROI^43^:

- Broca’s area – Speech production region.
- Left SFL – Associated with motor speech coordination.

#### 2.4.3 Statistical Comparison

Cortical Thickness changes were assessed using a mixed-effect ANOVA comparison. To strengthen the analysis, a reference group was added, selected from the Michael J. Fox Foundation’s Parkinson’s Progression Markers Initiative (PPMI) dataset^48^. This group was matched for age, gender, and Hoehn and Yahr (H&Y) scores to the four participants in the dance intervention group, providing a baseline for comparison and enhancing the interpretation of observed changes^a^. The reference group demographics and their matched subjects are demonstrated in TABLE 2.

**TABLE 2.** PPMI Reference Group Demographics.

| PPMI Subject ID | Matched Subject | Age<br>Range | Gender | H & Y |
| --- | --- | --- | --- | --- |
| 115448 | 1 | 66-70 | M | 1 |
| 101482 | 2 | 56-60 | M | 2 |
| 3102 | 3 | 61-65 | M | 1 |
| 100017 | 4 | 56-60 | F | 1 |

### 2.5. DTI and DWI Analysis

#### 2.5.1. DWI Acquisition & Preprocessing

A subset of 4 PwPD participants underwent DWI scanning in January and April, yielding 8 scans total. Data were processed using MRtrix3^49^, including: denoising (dwidenoise), motion and Eddy current correction (dwifslpreproc), bias field correction (dwibiascorrect), registration to standard space using the Human Connectome Map^47^, fiber orientation estimation using Constrained Spherical Deconvolution (CSD), and whole-brain probabilistic tractography (iFOD2).

#### 2.5.2. ROI-Based Connectivity Analysis

Tractography examined connectivity between Broca’s^47^ and the left SFL areas^47^.

10 million streamlines were initially generated, with ROI-based filtering used to isolate fibers connecting these regions.

#### 2.5.3. Statistical Comparison

White matter integrity was assessed using FA, MD, AD, and RD. Comparisons were performed using a paired t-test.

### 2.6. Statistical Analysis

Statistical analyses were conducted using SciPy library (1.14.1)^50^, within JupyterLab environment (version 4.2.5), which was accessed through ANACONDA Navigator (version 2.6.5) on a MacOS Sequoia 15.5 system. Statistical significance was set at *p* < 0.05 for all comparisons.

## 3. RESULTS

FIGURE 3 illustrates the F0SD trajectories for three PwPD participants (Subject IDs 1, 2, 3) over multiple sessions conducted from 2014 to 2019.

**FIGURE 3.**
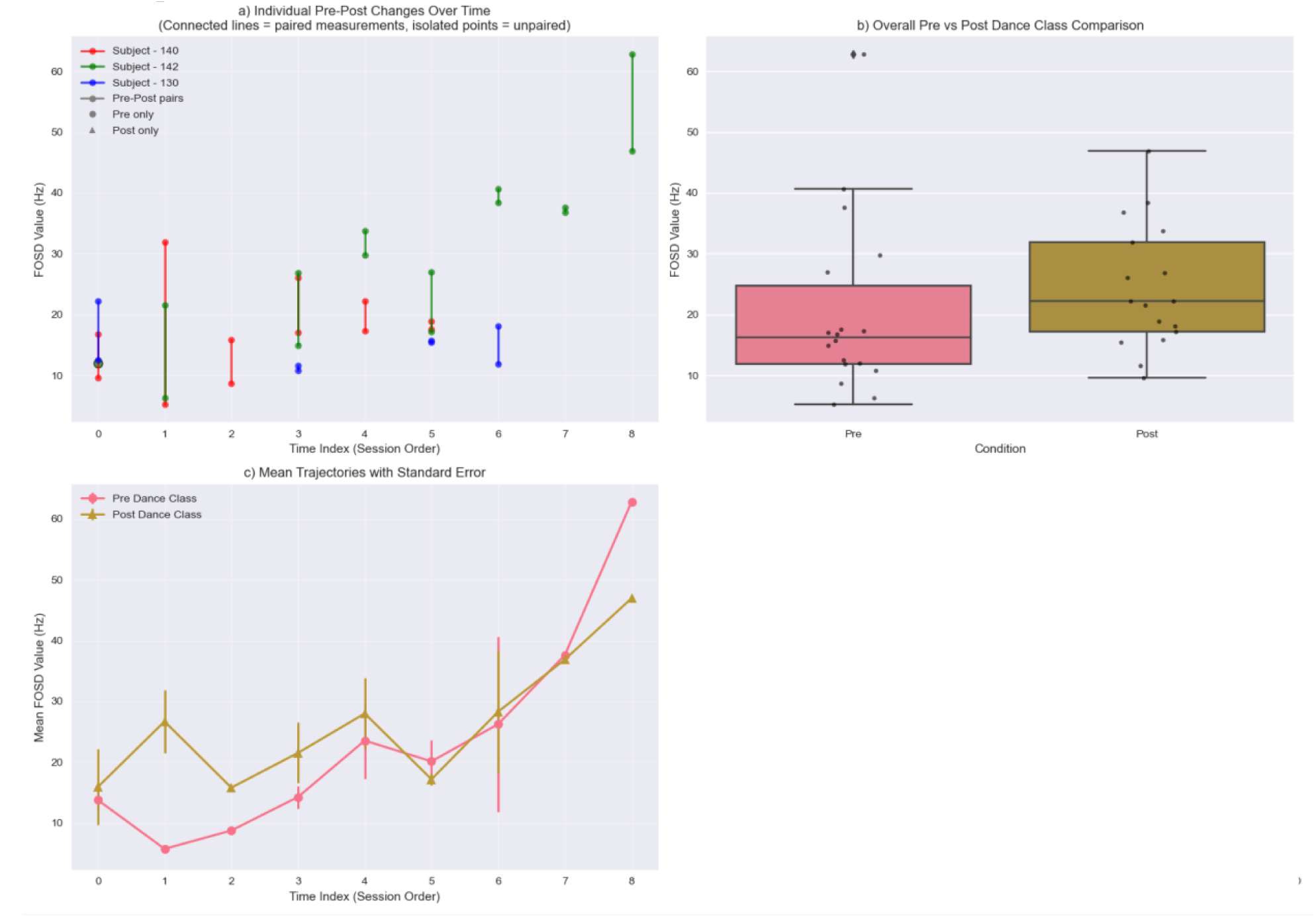
Fundamental frequency standard deviation (F0SD) in three people with Parkinson’s disease (PwPD) following dance training. a) individual pre- and post-dance values for F0SD in Hertz (Hz), b) overall comparison of pre- versus post-dance, c) mean F0SD changes across time with standard error. The effect of time was significant for F0SD changes in mixed-effect ANOVA model (*p* < 0.0001).

### 3.1 fMRI Results

fMRI analyses examined BOLD signal changes in the motor cortex (inferior frontal gyrus) and the SMA across four time points (September, December, January, April).

FIGURE 4 shows a significant decrease from September to January (*p* = 0.021), and the difference between September and December signals was marginally significant (*p* = 0.055) for the motor cortex. No statistically significant differences were observed between subsequent time points.

**FIGURE 4.**
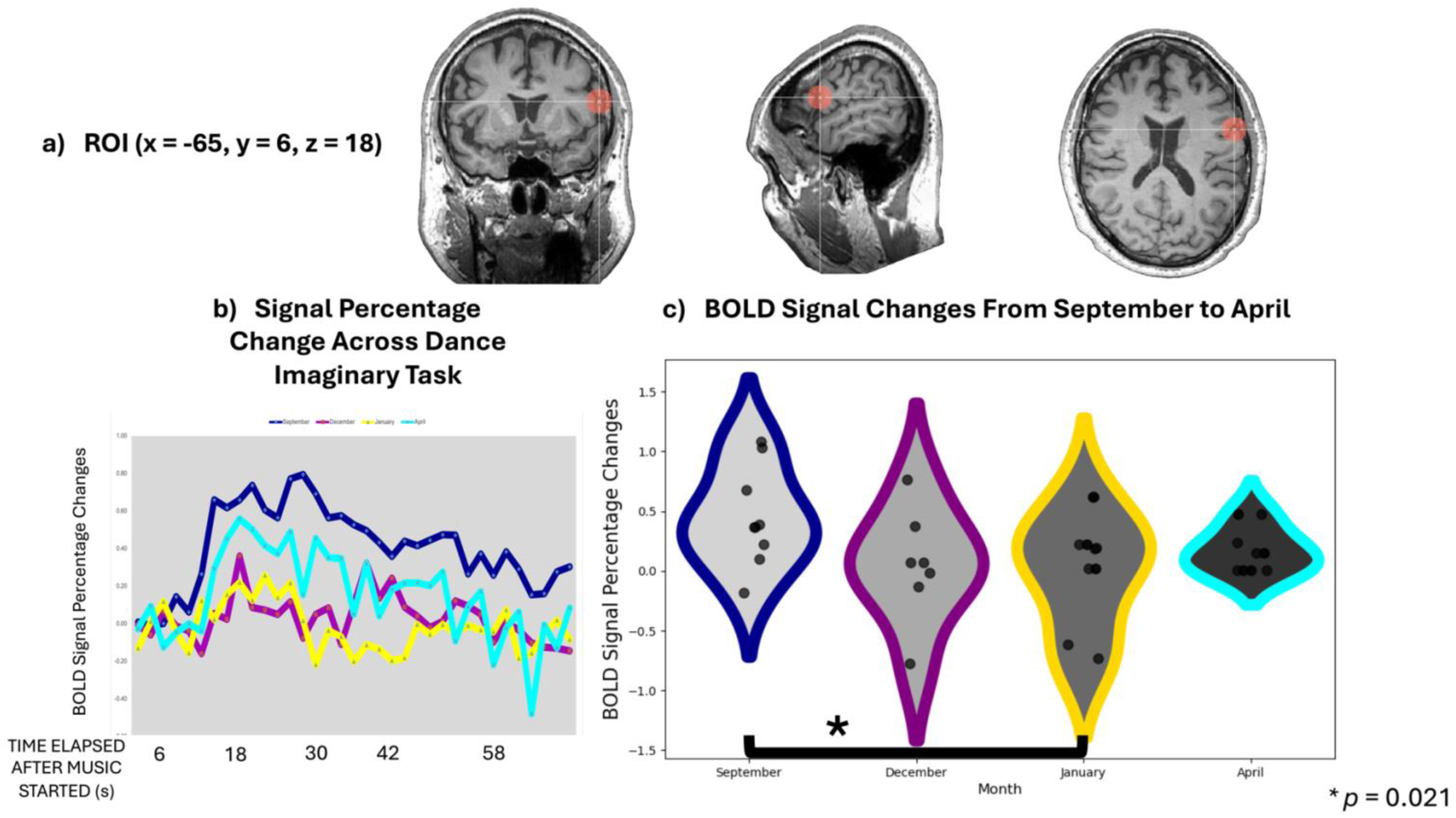
a) Inferior frontal gyrus Talairach coordinates. b) Signal percentage changes in inferior frontal gyrus during imaginary task in September, December, January, and April. c) BOLD signal percentage changes in the inferior frontal gyrus from September to April.

TABLE 3 presents the results of the mixed linear model regression for the BOLD signal changes in the inferior frontal gyrus.

**TABLE 3.**
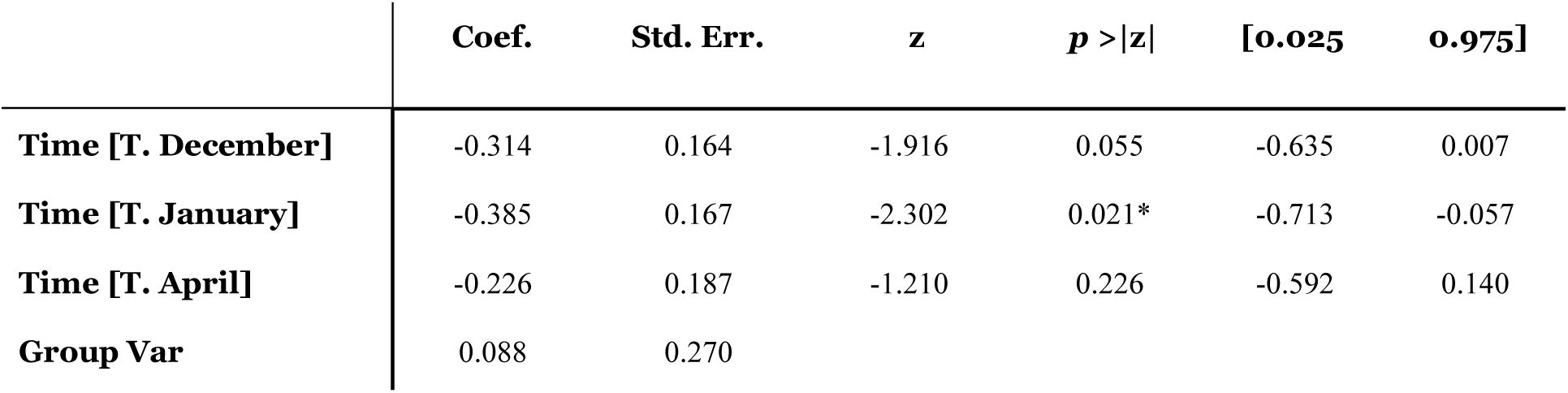
Mixed Linear Model Regression Results (Inferior Frontal Gyrus)

FIGURE 5 shows a significant increase from September to January (*p* = 0.010) in the SMA. No statistically significant differences were observed between subsequent time points.

**FIGURE 5.**
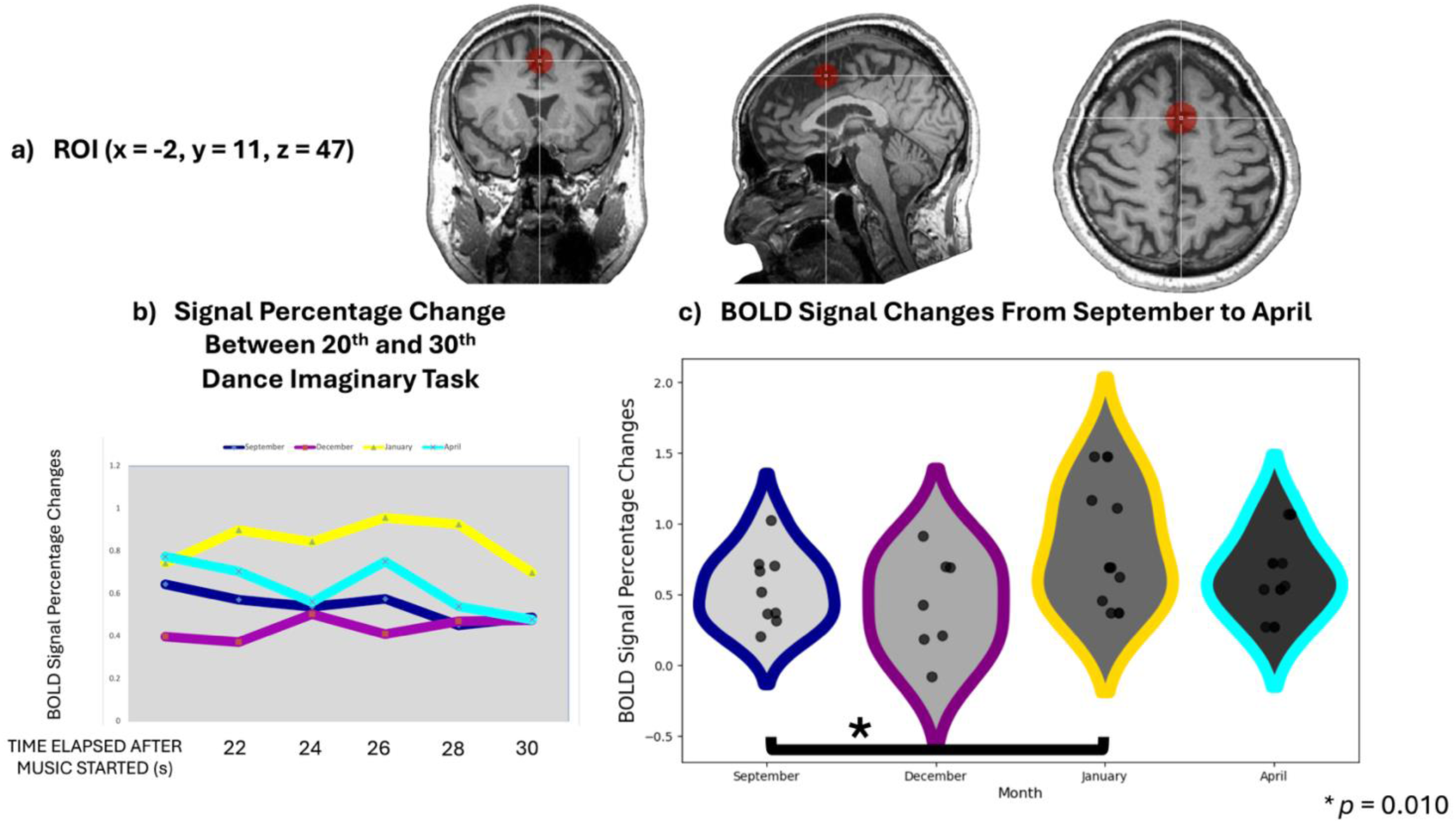
a) Supplementary motor area Talairach coordinates. b) Signal percentage changes in supplementary motor area during the vocal part of imaginary task in September, December, January, and April. c) BOLD signal percentage changes in the supplementary motor area from September to April.

TABLE 4 presents the results of the mixed linear model regression for the BOLD signal changes in the SMA.

**TABLE 4.** Mixed Linear Model Regression Results (SMA)

| | Coef. | Std. Err. | z | $p > \vert z\vert $ | [0.025 | 0.975] |
| --- | --- | --- | --- | --- | --- | --- |
| Time [T. December] | -0.101 | 0.129 | -0.782 | 0.434 | -0.354 | 0.152 |
| Time [T. January] | 0.343 | 0.133 | 2.547 | 0.010* | 0.082 | 0.605 |
| Time [T. April] | 0.150 | 0.151 | 0.992 | 0.321 | -0.146 | 0.446 |
| Group Var | 0.042 | 0.143 |  |  |  |  |

### 3.2 Cortical Thickness Results

Cortical thickness changes were analyzed for Broca’s and the left SFL areas across the four-time points. Violin plots (**FIGURE 6a & b**) illustrate cortical thickness distributions for each time point. Broca’s area showed no statistically significant changes in cortical thickness across time points (*p* > 0.05). In contrast, the left SFL area showed significant increase from September to December scan (*p* = 0.049), and a decrease from December to April (*p* = 0.046).

**FIGURE 6.**
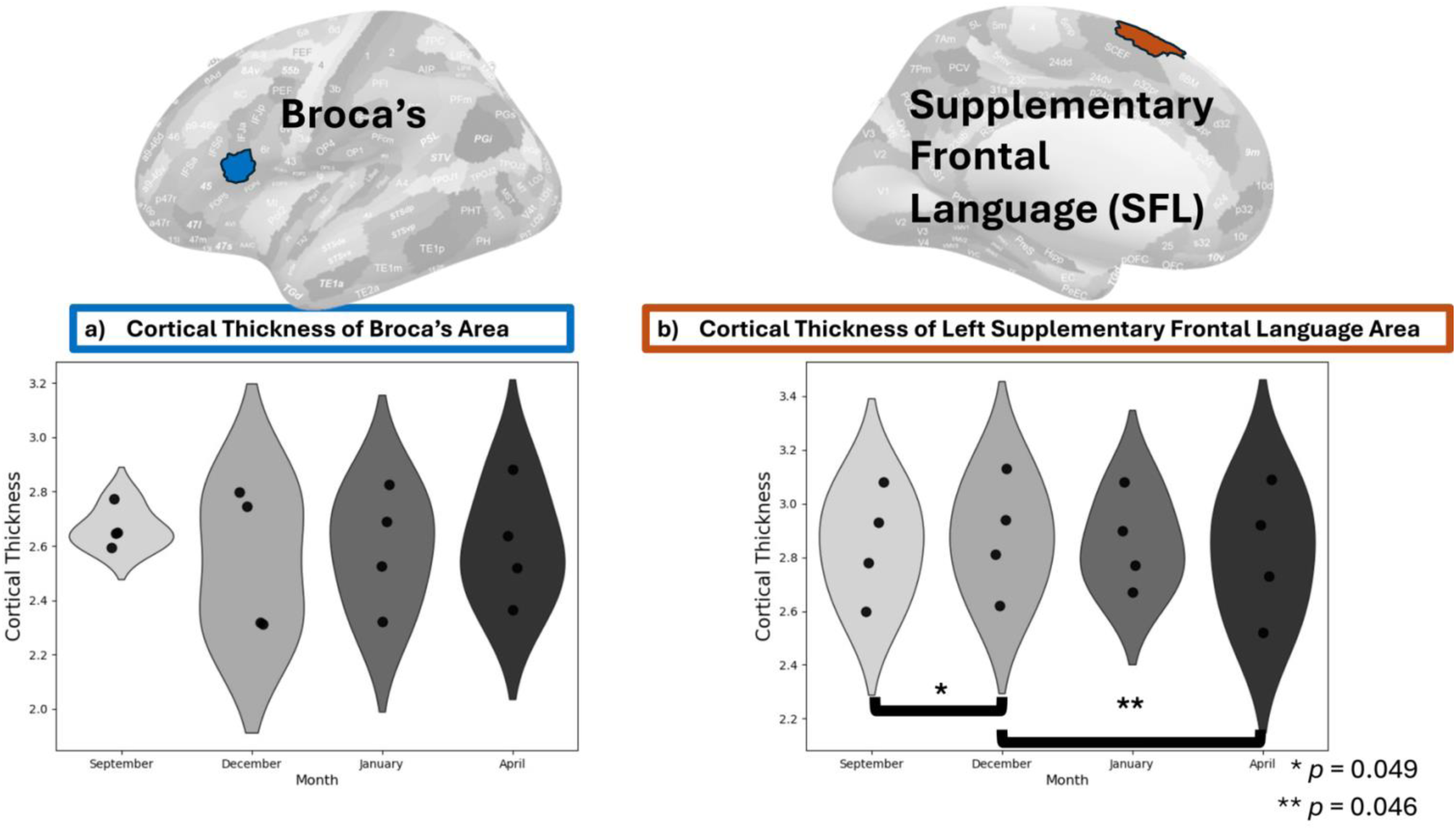
(a) Cortical thickness measurements of Broca’s area across the four-time points (September, December, January, and April). (b) Cortical thickness measurements of the left supplementary frontal language (SFL) area across the same time points.

The cortical thickness analysis in the PPMI reference group across a comparable 1-year time window revealed no significant changes in cortical thickness for either Broca’s area or the left SFL region during that period (FIGURE 7a & b).

**FIGURE 7.**
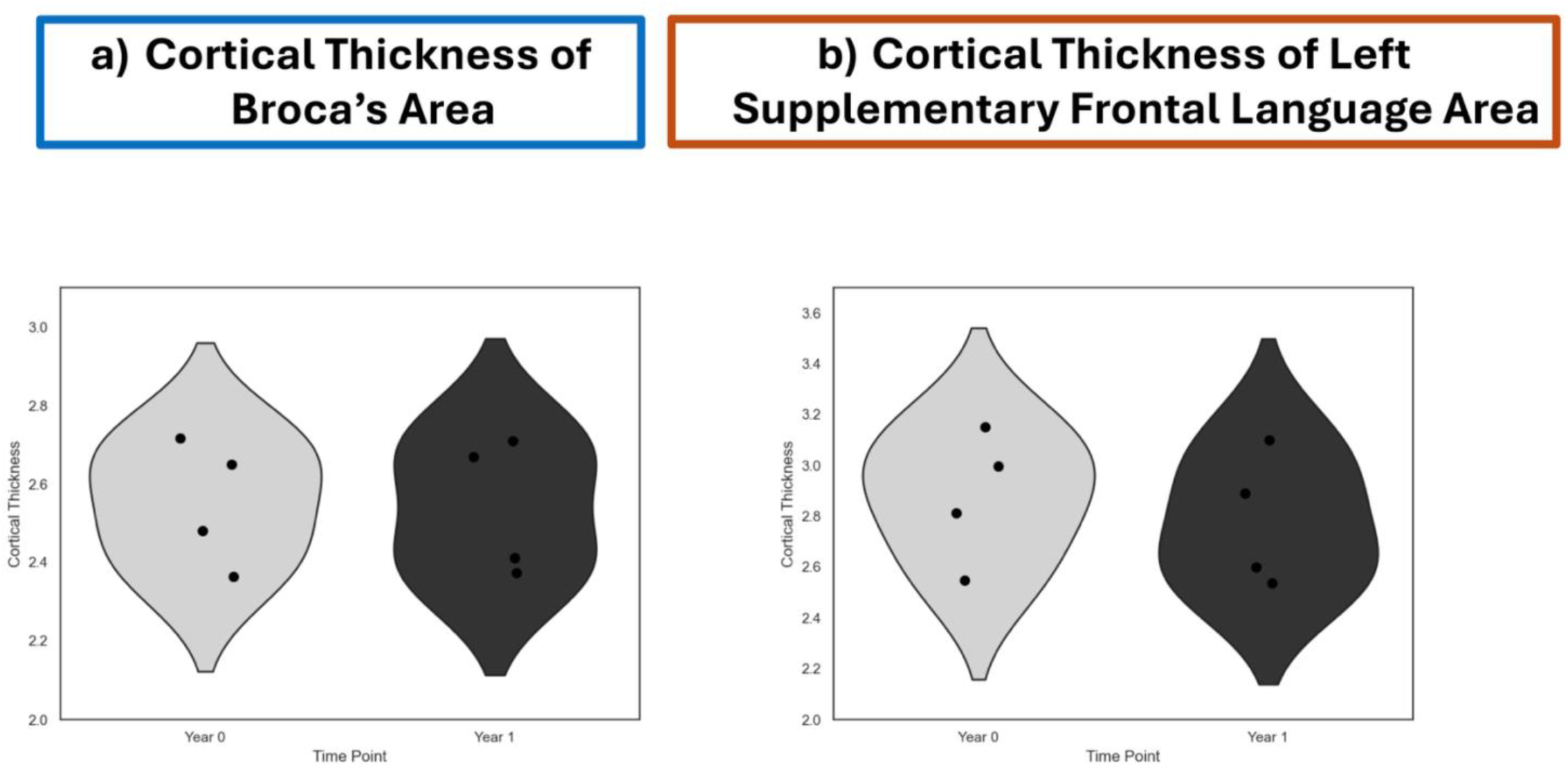
(a) Cortical thickness measurements of Broca’s area (a) and the left supplementary frontal language (SFL) (b) in the PPMI reference group across one year

TABLE 5 and TABLE 6 summarize the cortical thickness for both groups’ mean across time for Broca’s area and the left SFL, respectively.

**TABLE 5.** Cortical Thickness Means (± Standard Deviation) For Broca’s Area.

|  | TIME 1 | TIME 2 | TIME 3 | TIME 4 |
| --- | --- | --- | --- | --- |
| **DANCE GROUP** | $2.66675 \pm 0.076687$ | $2.5435 \pm 0.264167$ | $2.59075 \pm 0.217993$ | $2.60125 \pm 0.217972$ |
| **PPMI REFERENCE GROUP** | $2.5535 \pm 0.159884$ | - | - | $2.54125 \pm 0.17255$ |

**TABLE 6.**
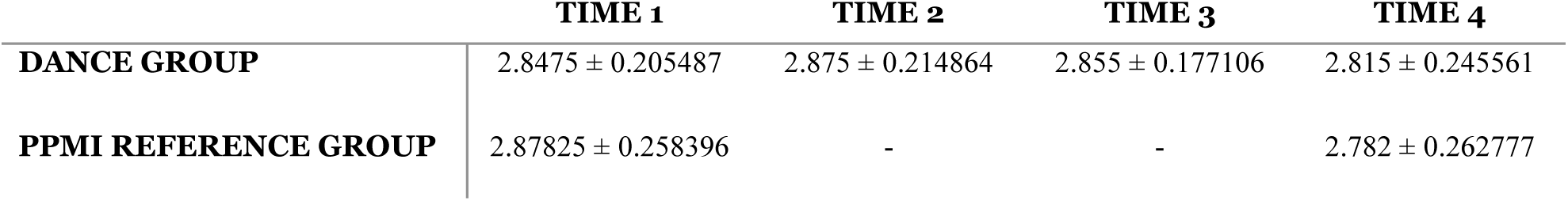
Cortical Thickness Means (± Standard Deviation) For Left Supplementary Frontal Language Area.

|  | TIME 1 | TIME 2 | TIME 3 | TIME 4 |
| --- | --- | --- | --- | --- |
| **DANCE GROUP** | $2.8475 \pm 0.205487$ | $2.875 \pm 0.214864$ | $2.855 \pm 0.177106$ | $2.815 \pm 0.245561$ |
| **PPMI REFERENCE GROUP** | $2.87825 \pm 0.258396$ | - | - | $2.782 \pm 0.262777$ |

### 3.3 DTI and DWI Results

DWI was performed in January and April to assess the integrity of white matter between Broca’s and the left SFL areas^b^. The tracks connecting Broca’s to the left SFL for each subject’s scan in January and April are visualized in FIGURE 8.

**FIGURE 8.**
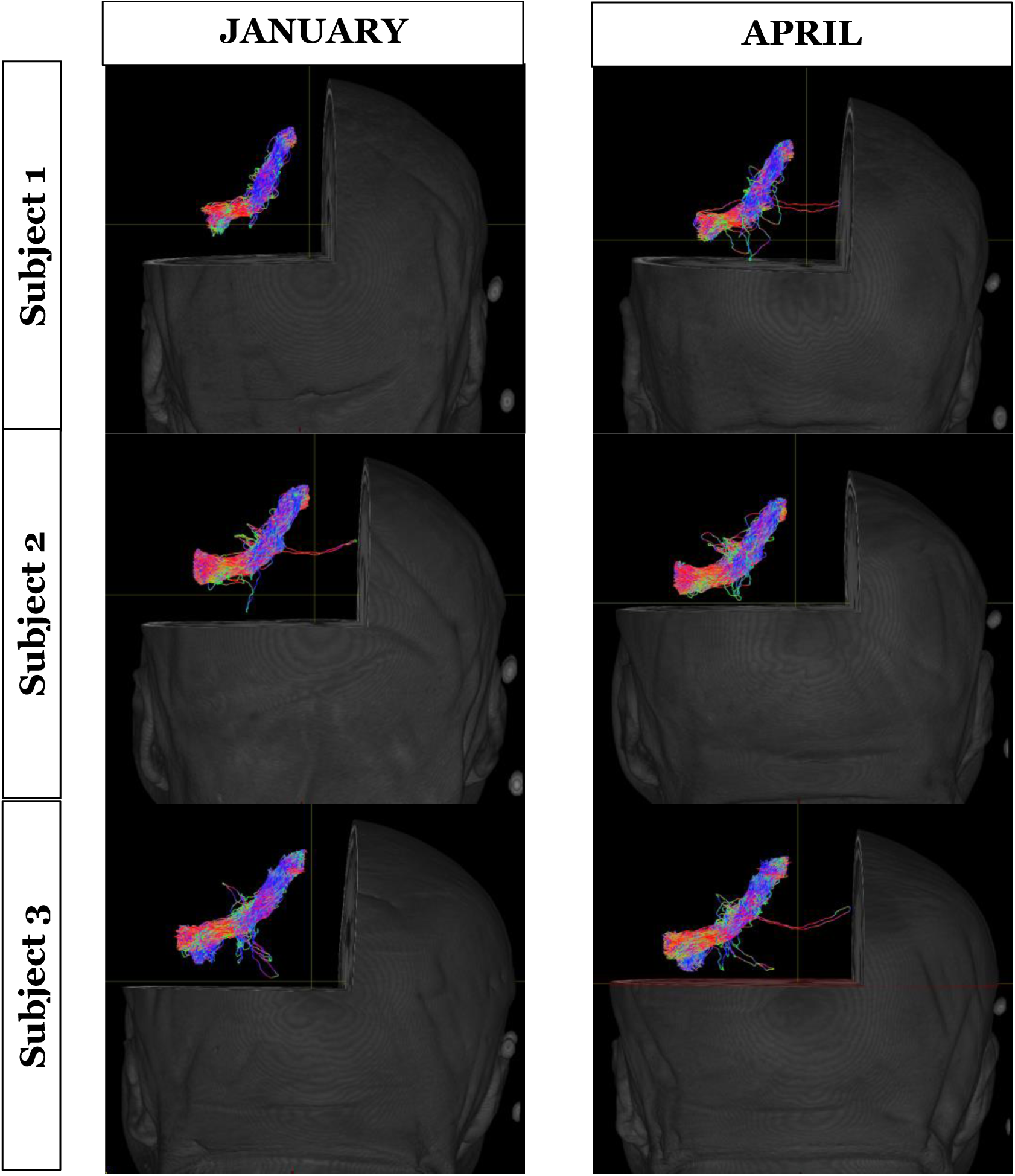
The tracks connecting Broca’s to the left supplementary frontal language area for each subject in January and April

The result indicated that FA significantly increased from January to April (*p* = 0.0247) (FIGURE 9a). AD showed a marginal increase (*p* = 0.056) (FIGURE 9b). RD and MD did not express significant changes (*p* > 0.05).

**FIGURE 9.**
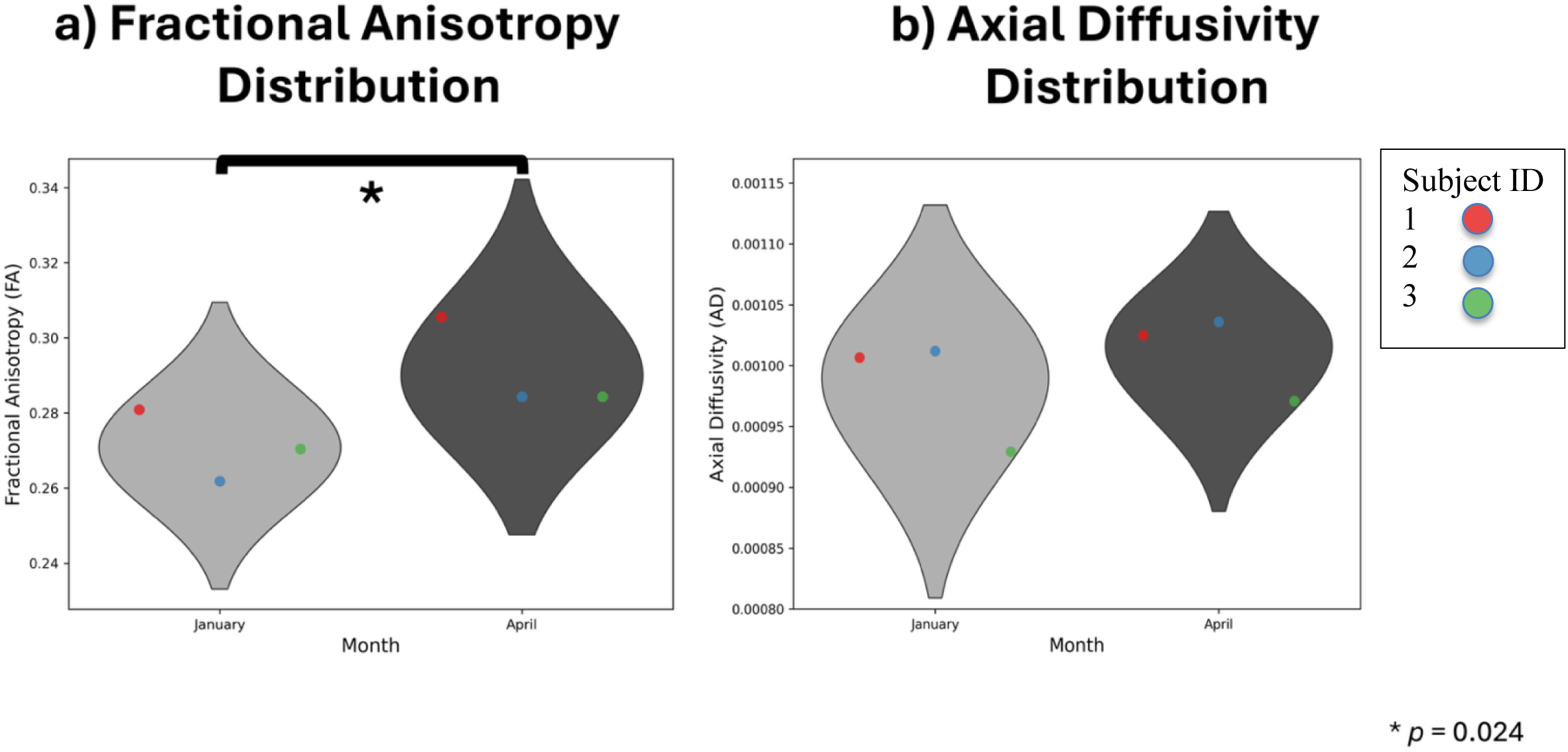
(a) Fractional anisotropy distribution and (b) Axial diffusivity distribution for tracks between the Broca area and the left supplementary frontal language (SFL) in January and April scans

## 4. DISCUSSION AND CONCLUSION

This study investigated the neuroplastic effects of dance training in PwPD by examining functional brain activity, cortical structure, and white matter connectivity changes across an 8- month period. Specifically, we utilized a multi-modal neuroimaging approach combining fMRI, cortical thickness analysis, and DWI. Our findings demonstrate significant neuroplastic adaptations in motor-related brain regions and white matter pathways, providing novel insights into the neural mechanisms underlying dance-induced improvements in PD.

### 4.1. Functional Changes in the Motor Cortex and the SMA

The analysis of F0SD trajectories in three participants (from 2014 to 2019) who completed both the neuroimaging study, and the subsequent longitudinal follow-up provides compelling evidence for the clinical relevance of the observed neuroplastic changes. The significant improvements in F0SD over time (*p* < 0.0001) demonstrate that the early neuroplastic adaptations detected in our 8-month imaging study were associated with sustained behavioral benefits that persisted for years.

The fMRI analysis revealed two distinct patterns of functional change over the course of the dance intervention. In the motor cortex (inferior frontal gyrus), there was a significant decrease in BOLD signal activity from baseline (September) to January. This reduction may reflect increased neural efficiency due to dance training. Motor imagery tasks typically activate motor cortical areas, and the observed decrease in BOLD signal over repeated sessions suggests a reduced neural demand for executing mental rehearsals of dance choreography, consistent with the motor learning literature^51–53^. Although no further significant differences were found beyond January, the stabilization of BOLD signals suggests that neuroplastic adaptations may reach a plateau with sustained training. These findings align with previous studies demonstrating that the initial phases of skill acquisition elicit greater cortical activation, which subsequently declines as performance improves, and motor processes become automated^54–56^.

In contrast, the SMA exhibited a significant increase in BOLD signal activity from September to January. The SMA plays a crucial role in motor planning, movement initiation, and internal generation of movement sequences, which are highly engaged during mental rehearsal tasks. The observed increase in SMA activation may reflect enhanced engagement of motor planning networks as participants became more adept at the cognitive and motor imagery aspects of dance rehearsal. Similar to the motor cortex, no further significant changes in SMA activation were observed beyond January, suggesting that functional adaptations may stabilize with continued training.

Overall, these findings indicate that dance training is associated with region-specific functional plasticity in PwPD, characterized by decreased activation in primary motor regions and increased engagement of higher-order motor planning areas during dance imagery tasks.

### 4.2. White Matter Integrity Between Broca’s Area and Left SFL

The significant increase in FA between Broca’s area and the left SFL area (*p* = 0.024) provides compelling evidence for structural neuroplasticity in pathways critical for speech-motor coordination. This finding is particularly relevant given the speech improvements documented in our F0SD analysis.

Structural Basis for Functional Improvements: The enhanced white matter integrity between Broca’s area and the left SFL area likely provides the structural foundation for the improved speech-motor coordination reflected in the F0SD improvements. Broca’s area is essential for speech production and motor speech planning, while the SFL area contributes to the coordination of complex motor sequences, including those required for prosodic speech control. Strengthened connectivity between these regions would facilitate more efficient communication between speech planning and motor execution networks, directly supporting the enhanced prosodic variability observed in our participants.

Myelin Plasticity and Motor Learning: The marginal increase in AD (*p* = 0.056) alongside stable RD and MD suggests that the FA improvements reflect enhanced axonal coherence rather than gross tissue changes. These results are consistent with previous research indicating motor training can promote structural reorganization and enhanced connectivity in white matter pathways involved in motor control and speech-motor coordination^39,57^. This pattern is consistent with myelin plasticity, where repeated activation of speech-motor circuits during dance training may have promoted myelin remodeling to support faster and more reliable signal transmission.

Such microstructural changes would be particularly beneficial for the rapid, precise adjustments required for prosodic speech control, as evidenced by the F0SD improvements.

### 4.3. Cortical Thickness in Language-Related Regions

The dynamic changes in left SFL cortical thickness - initial increase from September to December (*p* = 0.049) followed by decrease from December to April (*p* = 0.046) - while Broca’s area remained stable, provide insights into the temporal dynamics of structural plasticity during motor-speech training.

The biphasic pattern in SFL thickness may reflect an adaptive remodeling process where initial training demands lead to cortical expansion, followed by pruning and optimization as skills become more automated. This interpretation is consistent with motor learning literature showing that cortical thickness changes often follow inverted-U patterns during skill acquisition^58^. The fact that this dynamic process occurred in the SFL area—a region critical for motor-speech coordination—while participants were simultaneously showing improved F0SD suggests that the cortical remodeling was functionally relevant to speech-motor integration.

Specificity of Plasticity: The selective changes in the left SFL area, contrasted with stability in Broca’s area, suggest that dance training may particularly engage higher-order motor coordination processes rather than basic speech production mechanisms. This specificity may explain why dance training appears particularly effective for prosodic aspects of speech (as reflected in F0SD improvements) rather than basic articulatory functions, as prosody requires the complex temporal coordination of multiple motor systems that the SFL area helps orchestrate.

### 4.4. Limitations

While this study provides valuable insights into the neuroplastic effects of dance training in PwPD, several methodological limitations should be acknowledged. First, the study did not account for individual differences in disease duration, severity, and the age onset of disease which may have influenced the variability in observed neuroplastic changes. Additionally, factors such as medication status (ON versus OFF state) and individual responsiveness to dance training may have affected the results, potentially introducing variability in functional and structural brain adaptations. Another significant limitation is the absence of a control group; without a non-dance PwPD or healthy control group for fMRI and DWI, it is difficult to attribute the observed changes solely to dance training, as other factors such as natural disease progression or external lifestyle influences may have played a role. The limited sample size may have further reduced the statistical power necessary to detect subtle changes in cortical thickness or white matter connectivity. While significant changes in BOLD activity and FA values were observed, a larger sample could provide greater reliability and generalizability of the findings.

Moreover, participants may have exhibited differences in dance experience, movement ability, cognitive function, and adherence to the intervention, all of which could contribute to variability in neuroplastic adaptations. The heterogeneity of PD progression among participants might have also influenced the results, underscoring the need for future studies with more controlled participant selection criteria. Finally, the observational, non-randomized design of this study limits the ability to establish causality between dance training and neuroplastic changes. A randomized controlled trial (RCT) with matched control groups would provide stronger evidence for the specific contributions of dance training to neural adaptations in PD.

### 4.5. Implications and Future Directions

The present findings underscore the potential of dance-based interventions to induce neuroplastic adaptations in functional brain activity and white matter integrity among PwPD. While the overall cortical thickness changes were not significant, the observed functional and white matter connectivity modifications could be indicators of neuroplasticity. Given the limitations of the current study, including sample size and intervention duration, future research should explore these effects over longer periods and with larger cohorts. Integrating comprehensive behavioral assessments alongside neuroimaging could further elucidate the clinical relevance of these neural adaptations. Comparative studies examining different forms of physical and cognitive interventions could also provide insights into modality-specific effects on neuroplasticity and clinical outcomes in PD.

### 4.6. Conclusion

Our study provides the first comprehensive neuroimaging evidence that dance training may induce significant neuroplastic adaptations in speech-motor networks in PwPD. The multi-modal findings demonstrate enhanced neural efficiency in motor cortex, increased engagement of motor planning networks, and strengthened white matter connectivity between speech-motor regions. Critically, these neuroplastic changes are associated with sustained improvements in speech prosody, as evidenced by F0SD enhancements that persist for years following the initial intervention period.

The integration of neuroimaging findings with longitudinal behavioral data provides compelling evidence for the clinical relevance of dance-induced neuroplasticity. The temporal dynamics of these changes—with maximal neural adaptations occurring within the first 4-5 months of training but behavioral benefits continuing to accrue over years—suggest that dance training creates lasting structural and functional brain changes that support sustained therapeutic benefits.

These findings have important implications for the development of evidence-based, mechanism-informed interventions for speech and motor symptoms in PD. The demonstration that dance training can enhance white matter connectivity in speech-motor pathways while promoting neural efficiency suggests that integrative, multi-system approaches may be more effective than traditional, domain-specific therapies.

Future research should build upon these findings through larger, controlled studies that prospectively integrate neuroimaging and behavioral assessments, investigate the specific therapeutic components of dance training, and explore personalized intervention approaches. Such studies will be essential for translating these promising neurobiological findings into PD.

The convergence of neuroplastic adaptations and functional improvements demonstrated in this study supports dance training as a promising, evidence-informed intervention that warrants further investigation and clinical implementation for speech-motor rehabilitation in PD.

## ETHICAL CONSIDERATIONS

Written informed consent was obtained at each data collection time point throughout the 8- month data collection period, following an approved protocol from York University’s Ethics Board (2013-211 & 2017-296). The most recent Ethics approval (Certificate #: 2025-032) was obtained on February 21, 2025, from the York University Office of Research Ethics. All procedures adhered to the ethical standards of the institutional and national committees on human experimentation and complied with the Helsinki Declaration of 1975, as revised in 2000. Participants’ privacy and confidentiality were ensured through de-identification, and data were securely stored on password-protected external hard drives in the research laboratory at York University. Participants received CAD $25 per imaging session and CAD $50 reimbursement per visit to York University for imaging-related travel costs (CAD $1 ≈ USD $0.71).

## DATA AVAILABILITY STATEMENT

The study data is available from the corresponding author on reasonable request.

## ACKNOWLEDGMENTS

We extend our sincere gratitude to all members of our laboratory over the years who contributed to this project, as well as to the individuals at our various testing locations for their support. We are indebted to S. Robichaud, D. Rabinovich, R. Cohan, P. Dhami, S. Maguire, H. Tehrani, K. McDonald, and R. Andrew for invaluable assistance during all phases of the research program. We are especially grateful for the collaborative efforts and continued support from Canada’s National Ballet School (A. Seto and A. Powell), Dance with Parkinson’s Canada (S. Robichaud), Trinity St. Paul’s Church in Toronto, and the Dance for Parkinson’s program at the Mark Morris Dance Group (D. Leventhal).

Reference group data used in the preparation of this article were obtained from 2025-02-25 to 2025-02-28 from the Parkinson’s Progression Markers Initiative (PPMI) database (www.ppmi-info.org/access-dataspecimens/download-data), RRID: SCR_006431. For up-to-date information on the study, visit www.ppmi-info.org.

## SUPPLEMENTARY MATERIAL

### S.1 Expanded Cortical Thickness Analysis

In addition to the main analysis presented in the manuscript, cortical thickness measurements were also performed for all ten participants in the dance intervention group across the available time points (September, December, January, and April).

The main text focused on a subset of four participants who completed imaging at all four time points, ensuring consistency for longitudinal analysis. The remaining participants had missing scans at one or more time points, which precluded their inclusion in the primary longitudinal comparisons. However, to provide a more comprehensive view, we present here the results from all available data across the full cohort.

Cortical thickness values for Broca’s area and the left SFL area were extracted for each participant using FreeSurfer’s “recon-all” pipeline, following the same procedures described in the main text. Statistical comparisons were conducted using a linear mixed-effects model.

#### Broca’s area

- When considering all ten participants, no significant longitudinal changes in cortical thickness were observed in Broca’s area (*p* > 0.05).
- Individual variability was noted, with some participants exhibiting minor increases or decreases, but these did not reach statistical significance at the group level (**SUPPLEMENTARY FIGURE 1**).

**SUPPLEMENTARY FIGURE 1.**
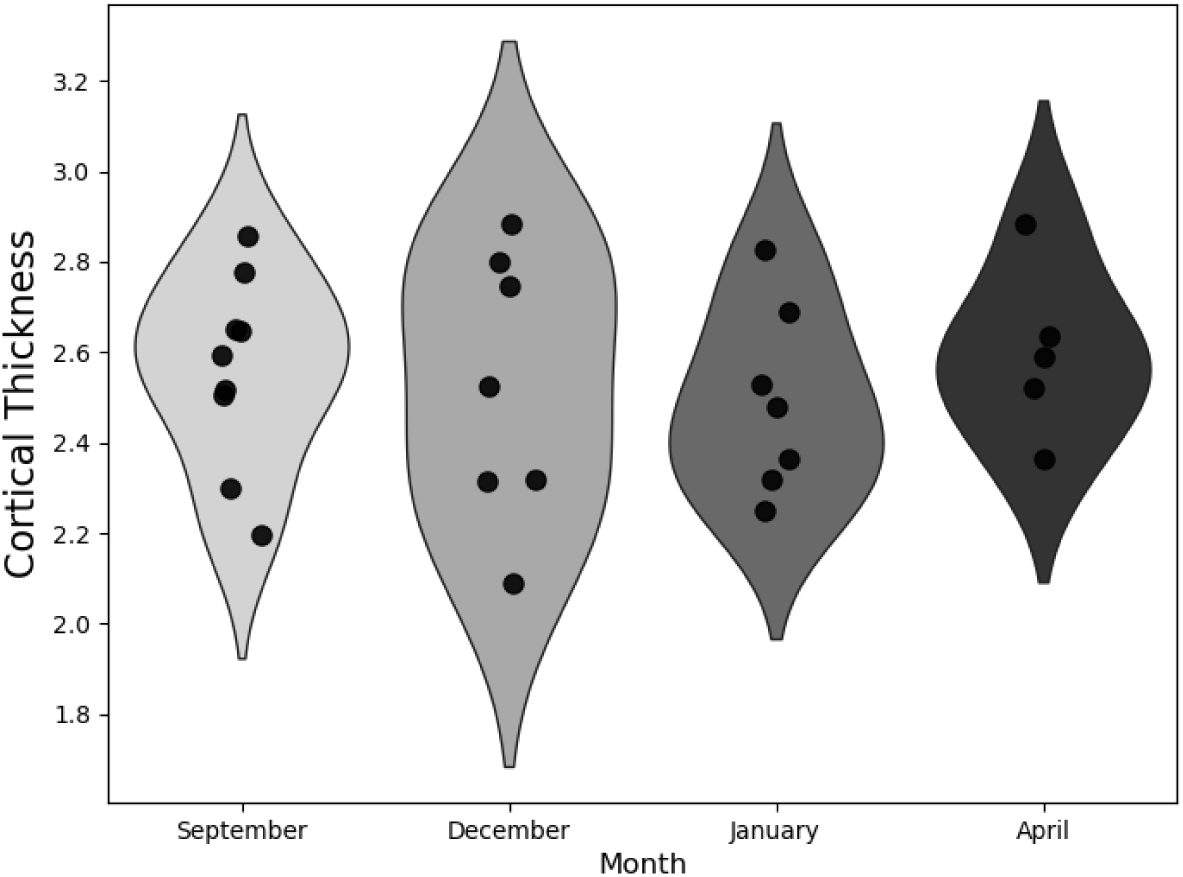
Violin plots showing the distribution of cortical thickness measurements across time points for Broca’s area (all available data from 10 subjects).

#### SFL area

- Similar to the subset analysis, the left SFL area showed a trend toward an initial increase in cortical thickness from September to December, followed by stabilization or slight decreases toward April (**SUPPLEMENTARY FIGURE 2**).
- However, these changes did not reach statistical significance when considering all ten subjects with available data (*p* > 0.05).

**SUPPLEMENTARY FIGURE 2.**
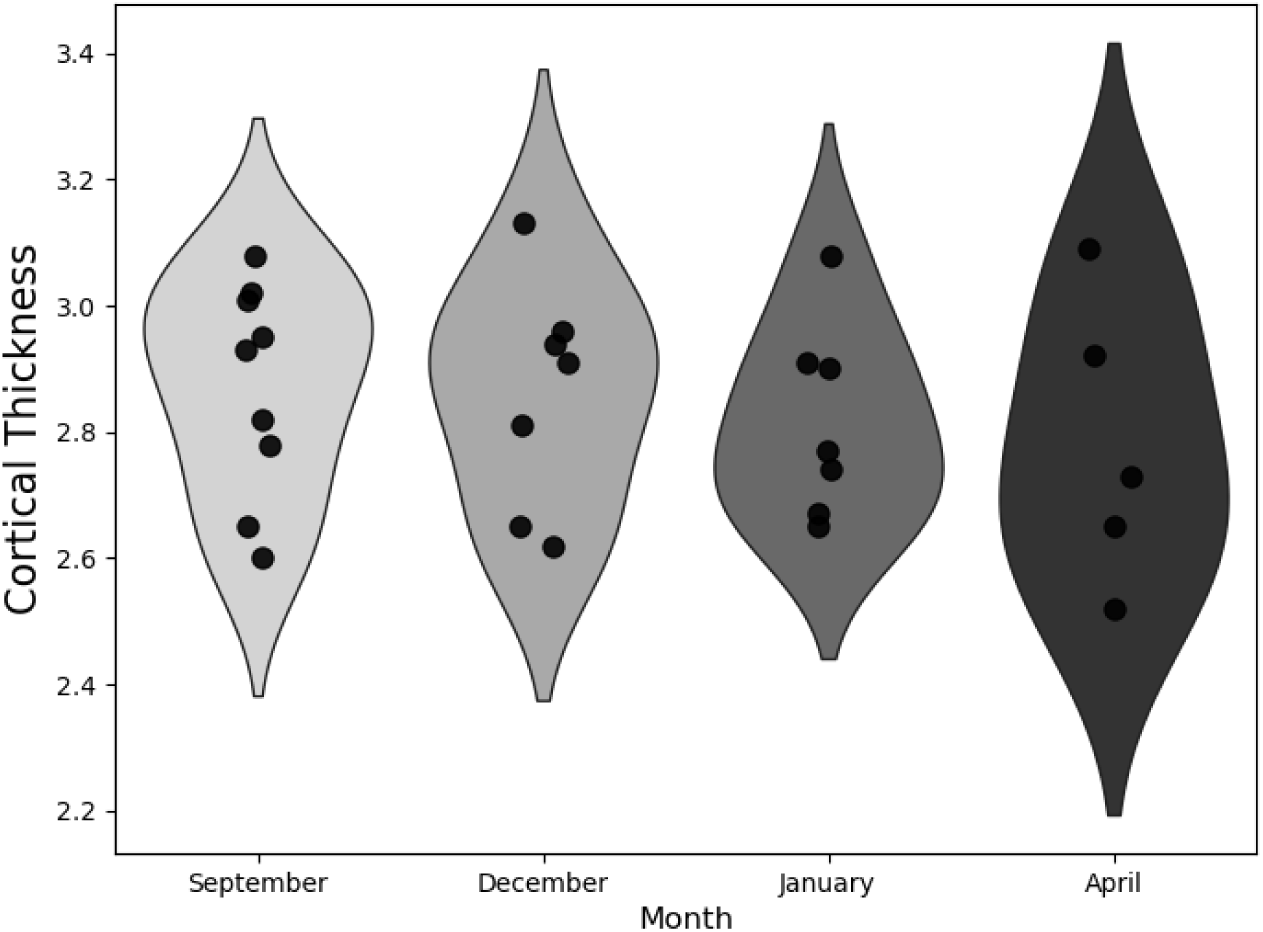
Violin plots showing the distribution of cortical thickness measurements across time points for the left supplementary frontal language (SFL) area (all available data from 10 subjects).

**SUPPLEMENTARY TABLE 1** represents the mean, median, standard deviation, minimum, and maximum values measured for the cortical thickness over Broca’s area and the left SFL area across 4 time points from the 10 subjects.

**SUPPLEMENTARY TABLE 1.** Descriptive Statistics of Cortical Thickness Values for Broca’s Area and Left Supplementary Frontal Language Area Across Time Points for All Participants.

| Area | TIME POINT | MEAN | MEDIAN | STD. DEV. | MIN | MAX |
| --- | --- | --- | --- | --- | --- | --- |
| Broca | September | 2.55988889 | 2.594 | 0.21087345 | 2.196 | 2.855 |
|  | December | 2.52428571 | 2.523 | 0.29757112 | 2.088 | 2.885 |
|  | January | 2.493 | 2.477 | 0.20734995 | 2.248 | 2.826 |
|  | April | 2.5988 | 2.589 | 0.18884835 | 2.366 | 2.883 |
| SFL | September | 2.86966667 | 2.931 | 0.16900148 | 2.599 | 3.077 |
|  | December | 2.85828571 | 2.908 | 0.18010248 | 2.623 | 3.13 |
|  | January | 2.81871429 | 2.774 | 0.15455497 | 2.649 | 3.084 |
|  | April | 2.7808 | 2.729 | 0.22436622 | 2.52 | 3.085 |

### S.2 Streamlines Between Broca’s and the Left SFL Areas

The mean, median, standard deviation, minimum and maximum length, and the number of tracks between Broca’s area and the left SFL are presented in **SUPPLEMENTARY TABLE 2**.

**SUPPLEMENTARY TABLE 2.** Descriptive Statistics of the Streamlines Between Broca’s and the Left Supplementary Frontal Language.

| **SUBJECT** | **TIME POINT** | **MEAN** | **MEDIAN** | **STD. DEV.** | **MIN** | **MAX** | **COUNT** |
| --- | --- | --- | --- | --- | --- | --- | --- |
| **140** | January | 73.6174 | 71.4445 | 10.9546 | 57.7505 | 149.444 | 408 |
|  | April | 73.2527 | 71.4309 | 11.7699 | 55.8497 | 196.196 | 533 |
| **142** | January | 78.0938 | 76.7318 | 9.83294 | 60.609 | 190.006 | 1224 |
|  | April | 77.9044 | 76.0529 | 11.3409 | 60.0604 | 188.076 | 876 |
| **130** | January | 89.2351 | 87.0606 | 13.5828 | 65.2059 | 188.583 | 1003 |
|  | April | 88.3505 | 86.6132 | 13.4954 | 64.5124 | 191.35 | 853 |

## Footnotes

a A supplementary analysis including all ten participants, regardless of missing time points, is provided in the **SUPPLEMENTARY MATERIAL** *section S.1*.

b The descriptive statistics for the tracks between the Broca’s and the left SFL are presented in **SUPPLEMENTARY MATERIAL** *section S.2*.

